# Interpretable AI for Accelerated Video-Based Surgical Skill Assessment: A Highlights-Reel Approach

**DOI:** 10.64898/2026.04.18.26351193

**Authors:** Marzie Lafouti, Liane S. Feldman, Amir Hooshiar

## Abstract

**Background:** Manual video-based evaluation of surgical skills can be time-consuming and delays trainee feedback. Artificial intelligence (AI) offers opportunities to automate aspects of assessment while maintaining clinician oversight. We developed an interpretable spatiotemporal model that classifies surgical expertise directly from endoscopic video in standardized training tasks and generates saliency-based “highlights reels” showing the most influential frames.

**Methods:** An RGB pipeline combining InceptionV3 for spatial feature extraction and a gated recurrent unit (GRU) for temporal modeling was trained on the JIGSAWS dataset. The model outputs novice, intermediate, or expert labels. A rolling-window, low-latency evaluation at 30 fps with a stride of 10 frames was used. A motion-augmented variant fused RGB with optical-flow features. Spatial and temporal saliency maps highlighted key decision-making regions.

**Results:** The RGB model achieved 95% accuracy (F1: 92% expert, 86% intermediate, 99% novice). Performance was strongest for novice and expert trials, while intermediate trials showed the lowest recall, consistent with greater ambiguity around the intermediate skill level. Saliency maps consistently emphasized tool–tissue interactions and peaked during technically demanding phases. The optical-flow variant underperformed, approximately 38% accuracy, which may reflect sensitivity to global camera motion and other non-informative motion patterns.

**Conclusions:** This interpretable AI pipeline accurately classifies surgical skill while producing intuitive visual highlights. Future work will refine highlight thresholds and validate on laparoscopic inguinal hernia repair for realworld deployment.

## 1. Introduction

Objective assessment of technical skill is a cornerstone of competency-based surgical education and has been shown to correlate with patient outcomes, operative efficiency, and error rates [1, 2]. With the increasing availability of intraoperative and simulation-based surgical video, video-based assessment has emerged as a promising modality for evaluating technical performance, enabling standardized viewpoints, asynchronous review, and longitudinal tracking of skill development [3, 4]. Despite these advantages, translating surgical video into timely, scalable, and actionable assessment remains challenging in routine educational practice.

Traditional video-based assessment relies heavily on expert raters, requiring substantial protected time for video review, scoring, and reconciliation across evaluators [2, 5]. These constraints limit the frequency with which assessments can be performed and delay feedback to trainees, reducing educational impact [4]. As surgical training programs move toward competency-based frameworks that emphasize continuous evaluation, the limitations of expert-dependent full-length video review have become increasingly apparent [3].

Recent advances in computer vision and deep learning have enabled automated analysis of surgical video for tasks such as workflow recognition, gesture segmentation, and skill classification [6, 7]. Several studies have demonstrated that deep learning models can distinguish between levels of surgical expertise directly from video data, suggesting the potential for objective, scalable performance evaluation [8]. However, despite strong predictive performance in experimental settings, adoption of these systems in surgical education remains limited.

Two key barriers have been repeatedly identified: first, the impracticality of reviewing full-length surgical videos at scale, even when AI-generated predictions are available; and second, the limited interpretability of deep learning models, which makes it difficult for surgeons to understand, validate, and trust automated assessments [9, 7, 10]. Without transparent justification of predictions and meaningful reduction in review burden, AI-based assessment tools risk remaining research prototypes rather than clinically integrated solutions.

In this work, we propose an AI-driven framework for accelerated video-based surgical skills assessment through the generation of interpretable surgical highlights reels. Our approach combines spatiotemporal deep learning with spatial and temporal saliency analysis to classify surgical expertise level while explicitly identifying where and when critical performance-related evidence occurs within a procedure. Building on these temporal importance signals, we introduce a relative-importance thresholding strategy to generate shortened videos and systematically evaluate whether highlights reels can preserve assessment performance while substantially reducing video length.

### 1.1. Background

Historically, surgical skill assessment has relied on expert observation and subjective judgment, either in the operating room or through video review. To improve objectivity and reliability, structured assessment instruments were developed, most notably the Objective Structured Assessment of Technical Skills (OSATS) [11]. OSATS demonstrated that global rating scales provide reliable discrimination between levels of technical expertise and outperform task-specific checklists in many contexts. Subsequent studies have validated OSATS across surgical specialties and simulation environments, establishing structured assessment as a foundation for competency-based education [5]. However, these approaches remain dependent on trained evaluators and protected time, limiting scalability when applied to large volumes of video data.

For laparoscopic procedures, the Global Operative Assessment of Laparoscopic Skills (GOALS) was introduced to capture dimensions such as depth perception, bimanual dexterity, efficiency, tissue handling, and autonomy [12]. GOALS and related global rating instruments have demonstrated strong validity and inter-rater reliability and are widely used in simulation-based laparoscopic training.

Despite their strengths, global rating scales require careful calibration of raters and remain inherently subjective. When applied to full-length operative videos, they impose a significant time burden that limits their feasibility for frequent formative assessment. Video-based assessment offers advantages over live observation, including repeatability and standardized viewpoints, but it remains labor-intensive and expert-dependent. Faculty time constraints, delayed feedback, and inter-rater variability have been repeatedly cited as barriers to widespread adoption of video review in surgical training programs [2, 4]. These challenges are amplified as video repositories grow and as competency-based frameworks demand more frequent assessment. Therefore, there is a clear clinical need for assessment tools that are scalable, objective, and capable of providing timely feedback. Reviews of machine learning in surgical education highlight the promise of automated video analysis while emphasizing the gap between algorithmic performance and clinical integration [7]. To be clinically useful, AI-based systems must reduce the burden of manual review while maintaining transparency and educational relevance. Beyond a numerical score, educators need interpretable evidence to support feedback and remediation. An ideal assessment system would therefore reduce the amount of video requiring review while highlighting the key moments and regions that drive assessment decisions.

### 1.2. Related Work

Automated assessment of surgical skill has been an active area of research for over a decade, driven by the need to complement expert-based evaluation with objective, scalable methods. Early work in this domain predominantly relied on hand-crafted features derived from tool motion, kinematics, and trajectory smoothness, particularly in robot-assisted surgery where motion data are readily available. Benchmark datasets such as JIGSAWS were introduced to facilitate comparative evaluation of gesture recognition and skill assessment methods by providing synchronized video and kinematic recordings across standardized tasks [13]. While kinematic-based approaches can capture fine-grained aspects of technical execution, their dependence on robotic platforms or specialized sensors limits generalizability to conventional laparoscopic procedures, where video is often the only consistently available modality.

With the advent of deep learning, researchers increasingly shifted toward end-to-end video-based approaches that learn discriminative representations directly from surgical footage. Convolutional neural networks (CNNs) have been employed to extract spatial features related to instrument appearance and scene context, while temporal modeling has been achieved using recurrent neural networks such as long short-term memory (LSTM) and gated recurrent unit (GRU) architectures [8]. These spatiotemporal pipelines have demonstrated strong performance in classifying surgical expertise and distinguishing between novice and expert behaviors on benchmark datasets, suggesting that video-only analysis can capture meaningful skill-related information.

More recent work has explored alternative spatiotemporal modeling strategies, including three-dimensional convolutional neural networks that jointly encode spatial and temporal information, as well as hybrid architectures that combine two-dimensional CNNs with temporal aggregation modules [6]. Two-stream networks incorporating motion representations such as optical flow have also been investigated in surgical video analysis, motivated by their success in general action recognition. However, surgical video presents unique challenges, including camera motion, specular highlights, smoke, and variable motion magnitude that can introduce noise into motion-based representations and limit their effectiveness in some contexts.

Accurate spatiotemporal modeling often depends on robust foundational scene understanding. Recent advancements have demonstrated the efficacy of deep learning methodologies for real-time object and instrument tracking in surgical videos, such as the use of Siamese networks [14] and deep fusion models like MedSAM-Flow [15]. Beyond tracking, broader scene inference and tissue segmentation pipelines, including those guided by natural language, have been developed to contextualize laparoscopic procedures [16, 17]. Preliminary conceptual frameworks have also recently proposed using AI-based content inference to generate shortened surgical highlights [18, 19].

Despite these advances, existing work has largely treated interpretability as a post hoc visualization tool rather than as a mechanism to directly reduce the burden of video review. Few studies have explicitly leveraged temporal importance information to generate shortened representations of surgical procedures and quantitatively evaluated whether such highlights can preserve assessment performance. Consequently, the feasibility of accelerated, highlights-based surgical skill assessment and the trade-off between video compression and predictive accuracy remain insufficiently explored. These gaps motivate the present study, which investigates whether interpretable spatiotemporal AI can be used not only to classify surgical skill, but also to generate surgical highlights reels that enable accelerated yet reliable video-based assessment.

### 1.3. Objective and Contributions

The primary objective of this study was to develop and evaluate an interpretable intelligent framework for accelerated video-based surgical skill assessment through AI-generated surgical highlights reels.

The specific contributions of this study were:

1. Development of a spatiotemporal deep learning pipeline that classifies surgical expertise level (novice, intermediate, expert) from video while generating spatial and temporal saliency maps.
2. Introduction of a relative-importance thresholding strategy to generate surgical highlights reels based on temporal saliency.
3. Quantitative evaluation of the accuracy–compression trade-off, demonstrating the feasibility and limitations of highlights-based assessment compared with full-length video analysis.

## 2. Methods

### 2.1. Dataset

All experiments in this study were conducted on the publicly available JHU-ISI Gesture and Skill Assessment Working Set (JIGSAWS), a benchmark dataset designed for comparative research in surgical gesture analysis and skill assessment [13, 20]. JIGSAWS contains synchronized endoscopic video and robot kinematics recorded during three standardized robot-assisted surgical training tasks: suturing, needle passing, and knot tying, performed by surgeons with varying experience levels [13, 20]. For each trial, an expertise label (novice, intermediate, expert) is provided as part of the dataset metadata [13].

In this work, we used the endoscopic video modality (RGB) as the primary input. Frames were sampled at the native frame rate (30 frames per second), and each frame was resized to match the spatial encoder input requirements (see Section 2.2). Standard ImageNet normalization was applied for CNN feature extraction. Videos were processed per task and per trial to ensure no leakage across training and evaluation partitions.

### 2.2. Model architecture and variants

We designed a spatiotemporal deep learning pipeline that maps RGB video to an expertise label (novice/intermediate/expert) while producing spatial and temporal saliency signals for interpretability (Figure 1). We used InceptionV3 to extract per-frame feature embeddings and a GRU for temporal modeling. Each RGB frame **x**_*t*_ was mapped to a feature vector **f**_*t*_ ∈ ℝ^*d*^ (global-average pooled from the penultimate layer), yielding a sequence {**f**_1_, …, **f**_*T*_}. The GRU processed this sequence to produce a video-level representation, which was mapped to a three-class softmax distribution (novice/intermediate/expert).

**Figure 1:**
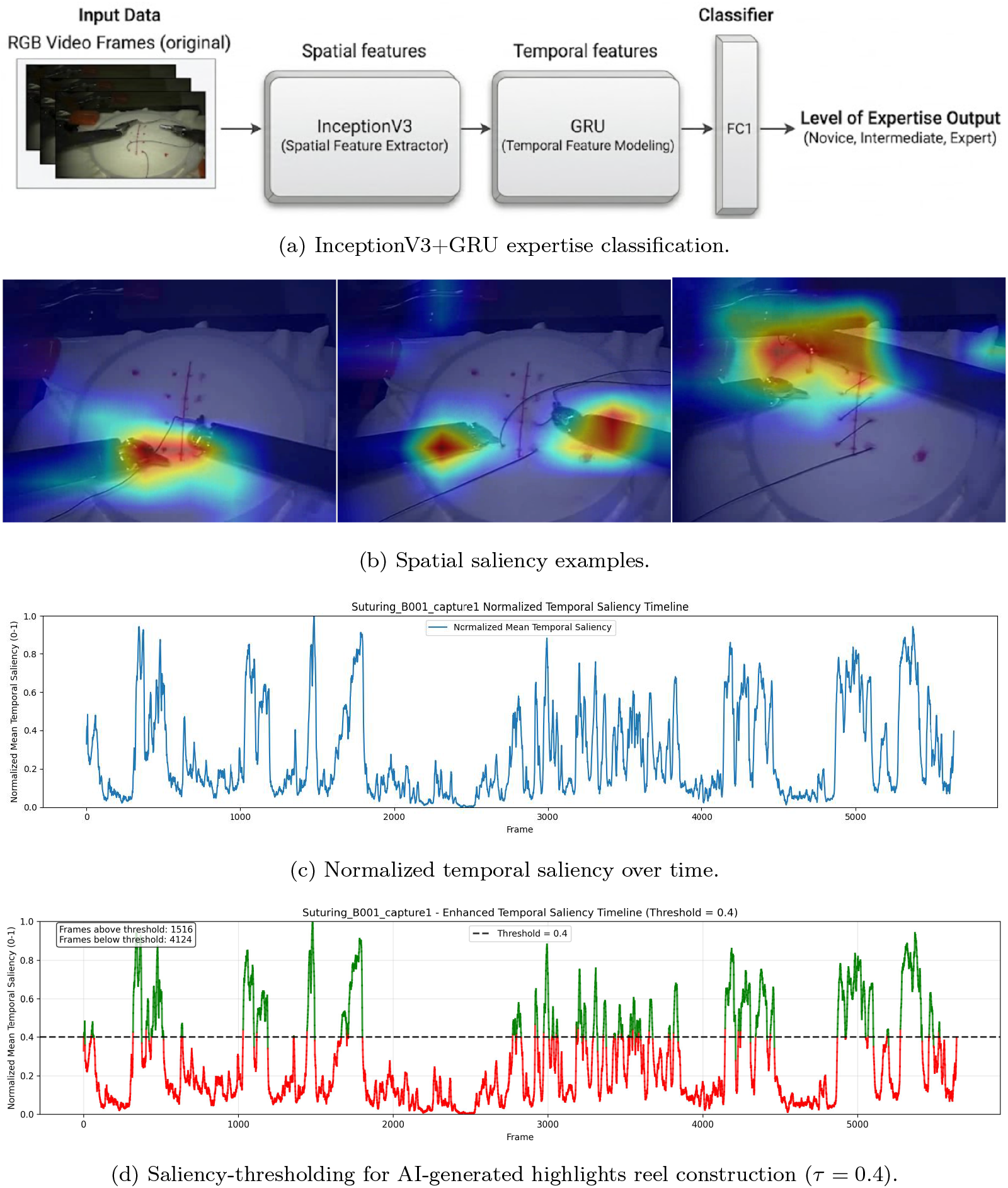
Overview of the proposed framework and interpretability outputs: (a) InceptionV3+GRU expertise classification, (b) spatial saliency examples, (c) normalized temporal saliency over time, and (d) saliency-thresholding for AI-generated highlights reel construction (example threshold *τ* = 0.4).

For low-latency evaluation and temporal interpretability, inference used overlapping windows of length *W* with a stride of 10 frames. Window-level class probabilities were averaged across windows to obtain a trial-level prediction.

#### 2.2.1. Motion-augmented variant with optical flow

To test whether explicit motion cues improve expertise classification, we implemented a motion-augmented variant inspired by two-stream video recognition concepts [21]. Dense optical flow was computed between consecutive frames using the TV-*L*^1^ method [22], producing flow fields that were encoded into flow images and processed through an analogous CNN feature extractor. Fusion was performed at the feature level (late fusion) by concatenating RGB and flow embeddings prior to temporal modeling.

#### 2.2.2. Saliency maps and spatiotemporal importance

To visualize which image regions most influenced predictions, we computed class-discriminative spatial saliency maps using Gradient-weighted Class Activation Mapping (Grad-CAM) applied to the last convolutional block of the spatial encoder [23]. For a target class *c*, Grad-CAM produces a coarse localization map 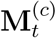 for each frame *t*, highlighting regions that contribute positively to the class logit. Example spatial saliency overlays are shown in Figure 1(b).

To estimate when the model relied most strongly on the input sequence, we computed a temporal saliency score per frame by measuring the sensitivity of the predicted class score to the corresponding frame-level embedding. Concretely, for the predicted class *c*^∗^, we computed the gradient magnitude

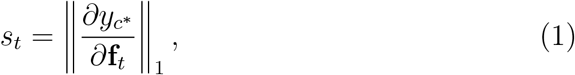

where *y*_*c*_∗ denotes the pre-softmax logit (or equivalently the probability after softmax, though logits are typically more stable). Frame-level scores were min-max normalized within each trial to obtain 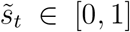. An example normalized temporal saliency profile is shown in Figure 1(c).

#### 2.2.3. AI-generated highlights reels via saliency thresholding

Using the normalized temporal saliency curve 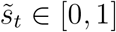, we generated an AI-based highlights reel for each trial by retaining only the most influential frames. As illustrated in Figure 1(d), a threshold *τ* was applied to the temporal saliency values: frames with 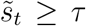 were kept (green), while frames with 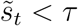 were discarded (red). Formally, the set of retained frame indices is

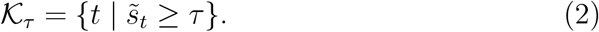

A shortened video was then constructed by concatenating the retained frames in their original temporal order. We evaluated multiple thresholds *τ* ∈ {0, 0.1, 0.2, …, 0.9}, where *τ* = 0 corresponds to the full video (no frame removal). For each *τ*, we re-ran inference on the shortened video and compared the resulting predictions with those obtained from the full-length video and with the ground-truth expertise labels.

To quantify the degree of shortening, we defined the retained-frame ratio (compression ratio) as

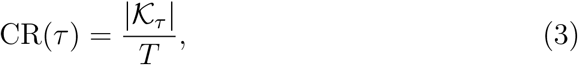

where *T* is the total number of frames in the original trial. Lower values of CR(*τ* ) indicate stronger compression (shorter highlights reels).

### 2.3. Training and evaluation

Model training was performed using task-stratified splits according to standard JIGSAWS evaluation protocols. In particular, we report results using a leave-one-super-trial-out (LOSO) cross-validation strategy, which is commonly used for JIGSAWS benchmarking and helps control for trial-level variability [8, 24]. Hyperparameters (optimizer, learning rate, batch size, and regularization) were selected on validation folds only.

Performance was quantified using overall accuracy, per-class precision, recall, and macro-averaged F1-score. Class-wise precision, recall, and F1-scores were used to characterize errors across skill levels. For highlights evaluation, we report the relationship between prediction performance and compression ratio across thresholds to identify feasible operating regions for accelerated assessment.

#### 2.3.1. Computing infrastructure

All experiments were conducted on a Lambda GPU workstation with an AMD Ryzen Threadripper PRO 5955WX CPU (16 cores), 512 GB RAM, and a NVIDIA RTX 6000 Ada Generation GPU.

## 3. Results

### 3.1. Overall expertise classification performance

On the JIGSAWS dataset, the RGB spatiotemporal model achieved an overall accuracy of 95% for three-class expertise classification including novice, intermediate, and expert (Table 1). Class-specific performance was strongest for novice and expert trials. Class-wise metrics showed the lowest recall for the intermediate class (Table 2), indicating that most ambiguity occurred around intermediate performance. The RGB model achieved high F1-scores across classes (Expert: 0.92, Intermediate: 0.86, Novice: 0.99). However, the RGB+optical-flow variant performed substantially worse (38.03% accuracy).

**Table 1:**
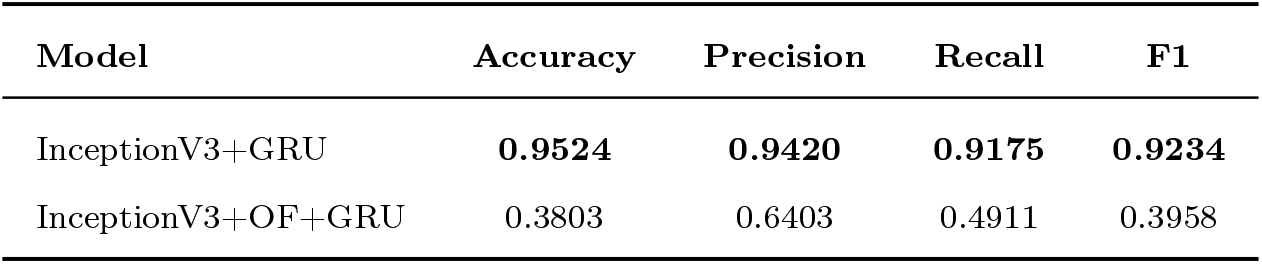
Overall metrics for expertise classification on JIGSAWS comparing RGB and RGB+optical flow models.

| Model | Accuracy | Precision | Recall | F1 |
| --- | --- | --- | --- | --- |
| InceptionV3+GRU | **0.9524** | **0.9420** | **0.9175** | **0.9234** |
| InceptionV3+OF+GRU | 0.3803 | 0.6403 | 0.4911 | 0.3958 |

**Table 2:** Class-wise metrics for expertise classification on JIGSAWS comparing RGB and RGB+optical flow models.

| Model | Class | Precision | Recall | F1 |
| --- | --- | --- | --- | --- |
| InceptionV3+GRU | Expert | 0.861 | 0.989 | 0.921 |
|  | Intermediate | **0.990** | 0.764 | 0.863 |
|  | Novice | 0.975 | **0.999** | **0.987** |
| InceptionV3+OF+GRU | Expert | 0.244 | 0.945 | 0.388 |
|  | Intermediate | 0.851 | 0.312 | 0.456 |
|  | Novice | 0.826 | 0.217 | 0.343 |

### 3.2. Interpretability: spatial and temporal saliency maps

Spatial saliency maps frequently highlighted regions around active instruments, particularly tool tips and tool-tissue interaction sites (Figure 2). Across tasks, salient regions often co-localized with instrument motion and suture/needle handling.

**Figure 2:**
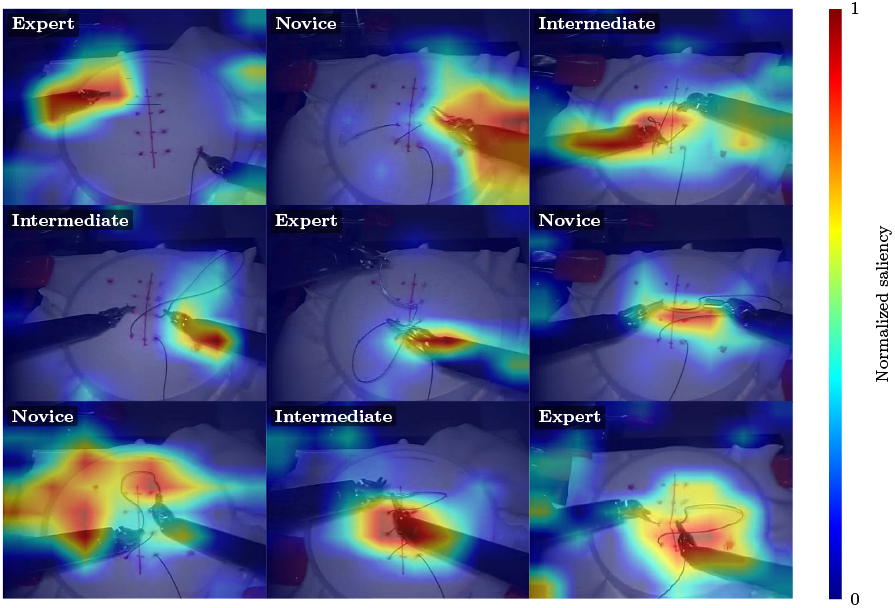
Representative spatial saliency maps overlaid on RGB frames. The predicted expertise class is shown in the top-left corner of each panel. The colorbar indicates normalized saliency intensity (0: low, 1: high); warmer colors denote regions contributing most to the predicted expertise class.

Temporal saliency was not uniform across trials. Higher saliency scores occurred in a subset of time points (Figure 1c), indicating that the model relied more strongly on specific segments of the procedure than on others.

### 3.3. Highlights-reel feasibility: effects of saliency thresholding

We next evaluated whether temporal saliency could be used to construct shortened videos (highlights reel) while preserving expertise classification performance. Increasing the saliency threshold *τ* reduced the proportion of retained frames and shortened the resulting highlights video, but also decreased predictive accuracy, demonstrating an explicit accuracy-compression trade-off. A representative example of the thresholding process is shown in Figure 3, illustrating how increasing *τ* progressively removes low-saliency frames and shortens the resulting highlights reel.

**Figure 3:**
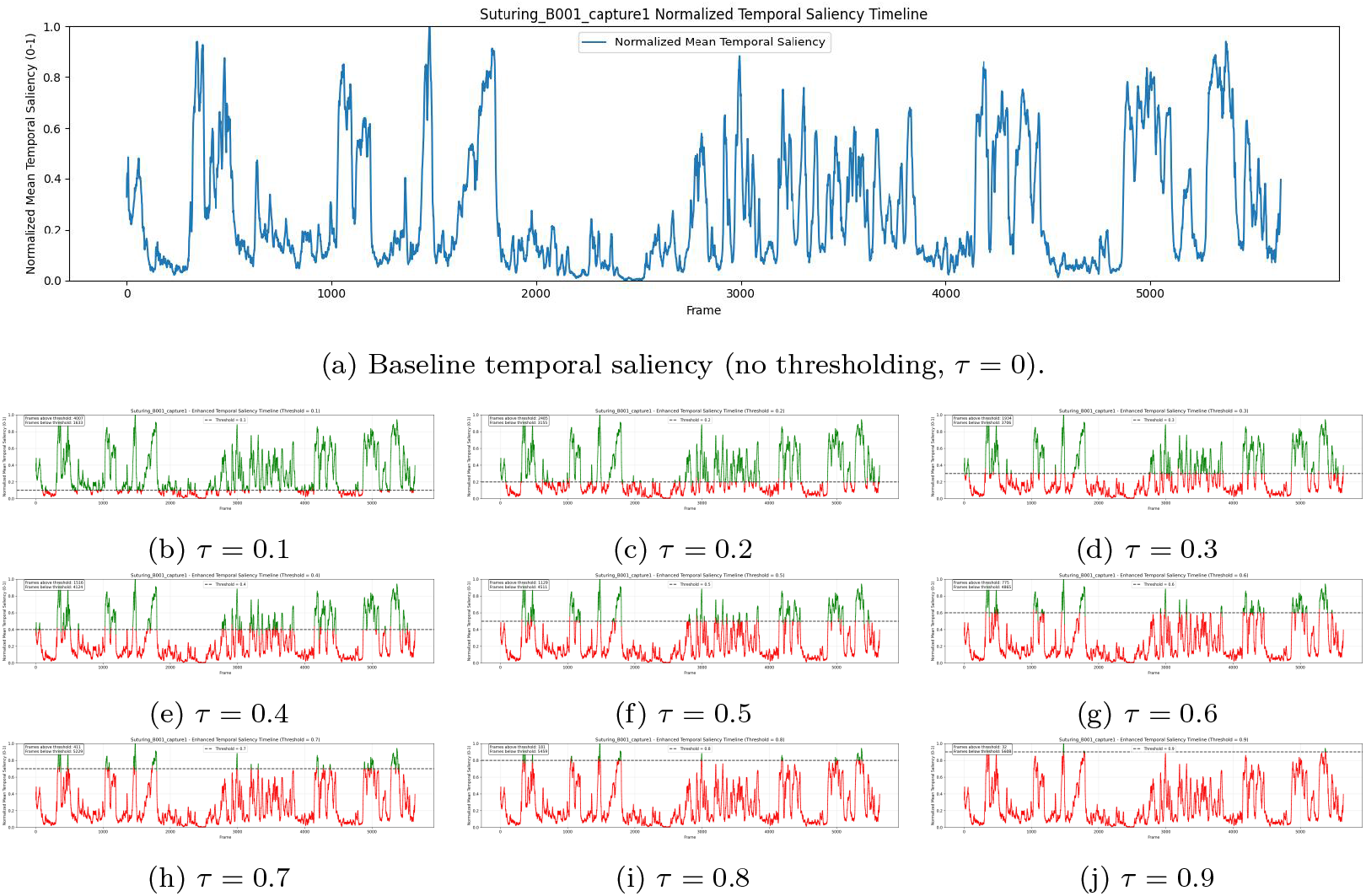
Effect of temporal-saliency thresholding on a representative suturing trial. Panel (a) shows the normalized temporal saliency curve (*τ* = 0). Panels (b)-(d) show example thresholds (*τ* = 0.1, 0.4, 0.8) illustrating retained (green) versus removed (red) frames.

Quantitative results across thresholds are reported in Table 3, with per-class trends in Table 4. Across thresholds, performance remained relatively stable at moderate compression levels, but degraded rapidly beyond a practical boundary. In particular, increasing *τ* from 0 to 0.4 reduced accuracy from 95% to 75%, and thresholds *τ >* 0.4 produced unreliable performance (Table 3). Figure 4 summarizes the relationship between threshold, retained-frame ratio, and accuracy.

**Table 3:** Highlights-reel performance across temporal-saliency thresholds.

| $\tau$ | Length | Retained (%) | Removed (%) | Acc. (%) | Prec. (%) |
| --- | --- | --- | --- | --- | --- |
| 0.0 | 00:03:07 | **100.0** | **0.0** | **95** | **94.20** |
| 0.1 | 00:02:13 | **71.1** | **28.9** | **92** | **91.32** |
| 0.2 | 00:01:22 | **43.9** | **56.1** | **88** | **89.07** |
| 0.3 | 00:01:04 | **34.2** | **65.8** | **81** | **80.44** |
| 0.4 | 00:00:50 | **26.7** | **73.3** | **75** | **79.58** |
| 0.5 | 00:00:37 | 19.8 | 80.2 | 52 | 58.63 |
| 0.6 | 00:00:25 | 13.4 | 86.6 | 43 | 48.21 |
| 0.7 | 00:00:13 | 7.0 | 93.0 | 49 | 49.08 |
| 0.8 | 00:00:06 | 3.2 | 96.8 | 41 | 42.48 |
| 0.9 | 00:00:01 | 0.53 | 99.47 | 48 | 51.25 |

**Table 4:** Per-class performance across temporal-saliency thresholds (Precision/Recall/F1).

| $\tau$ | Expert | | | Intermediate | | | Novice | | |
| --- | --- | --- | --- | --- | --- | --- | --- | --- | --- |
|  | P | R | F1 | P | R | F1 | P | R | F1 |
| 0.0 | 0.861 | 0.989 | 0.921 | 0.990 | 0.764 | 0.863 | 0.975 | 0.999 | 0.987 |
| 0.1 | 0.872 | 0.886 | 0.879 | 0.934 | 0.885 | 0.909 | 0.934 | 0.983 | 0.958 |
| 0.2 | 0.948 | 0.836 | 0.888 | 0.869 | 0.944 | 0.905 | 0.854 | 0.775 | 0.813 |
| 0.3 | 0.936 | 0.745 | 0.829 | 0.787 | 0.922 | 0.849 | 0.690 | 0.507 | 0.585 |
| 0.4 | 0.943 | 0.680 | 0.790 | 0.683 | 0.981 | 0.805 | 0.762 | 0.390 | 0.516 |
| 0.5 | 0.864 | 0.687 | 0.765 | 0.295 | 0.969 | 0.453 | 0.600 | 0.127 | 0.209 |
| 0.6 | 0.625 | 0.882 | 0.732 | 0.250 | 0.800 | 0.381 | 0.571 | 0.111 | 0.186 |
| 0.7 | 0.357 | 0.500 | 0.417 | 0.615 | 0.800 | 0.696 | 0.500 | 0.294 | 0.370 |
| 0.8 | 0.412 | 0.700 | 0.519 | 0.529 | 0.900 | 0.667 | 0.333 | 0.059 | 0.100 |
| 0.9 | 0.438 | 0.778 | 0.560 | 0.500 | 0.667 | 0.571 | 0.600 | 0.200 | 0.300 |

**Figure 4:**
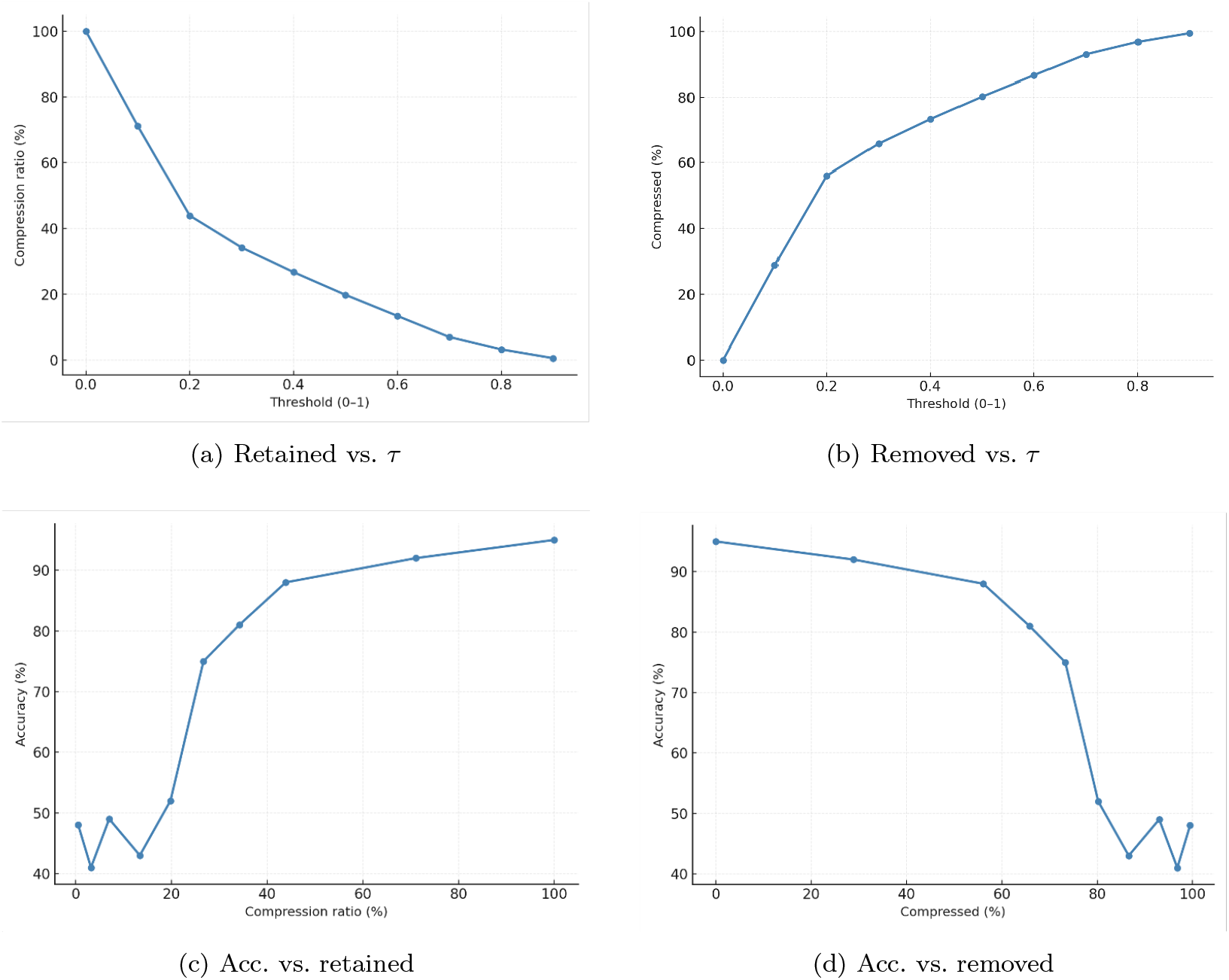
Accuracy–compression trade-off under saliency thresholding: retained/removed proportions vs. *τ* and accuracy vs. compression.

## 4. Discussion

This study demonstrates that an interpretable RGB spatiotemporal pipeline can classify surgical expertise on JIGSAWS with high accuracy while providing interpretable spatial and temporal saliency explanations. These findings support prior evidence that video-only deep learning can capture discriminative technical signatures of skill without relying on external sensors or kinematics [3, 8]. Importantly, the saliency outputs (Figures 1c and 2) add transparency to the predicted skill labels, addressing a key barrier to clinical adoption of AI decision support in surgery, namely, the need for interpretable, surgeon-verifiable model behavior [7].

Across representative trials, Grad-CAM maps emphasized tool tips, suture material, and tool-tissue interaction regions (Figure 2), which aligns with how expert raters conceptually judge economy of motion, tissue handling, and instrument control in validated assessment frameworks such as OSATS and GOALS [11, 12]. Temporal saliency further suggested that model evidence is concentrated in technically demanding phases rather than being uniformly distributed across the procedure (Figure 1c). This is consistent with the intuition that a subset of critical maneuvers (e.g., needle driving, knot formation, precise bimanual coordination) is disproportionately informative for global competency judgments. Beyond face validity, such spatiotemporal explanations may support more actionable feedback by linking performance scores to identifiable moments and regions, potentially complementing structured video debriefing approaches increasingly explored in surgical education [25].

A central contribution of this work is the use of temporal saliency to generate AI-driven highlights reels and quantitatively characterize the resulting accuracy-compression trade-off. As shown qualitatively in Figure 3 and quantitatively in Tables 3, 4 and Figure 4, increasing the threshold *τ* progressively removes low-saliency frames, shortening the video but degrading classification performance beyond moderate compression. If a conservative operating point is required (e.g., ≥ 90% accuracy in this benchmark setting), the results suggest using mild thresholding (approximately *τ* ≤ 0.1), whereas more aggressive thresholding (*τ* ≥ 0.4) substantially reduced reliability. In our experiments, performance remained acceptable at mild compression (e.g., *τ* = 0.1; ∼ 29% removed; 92% accuracy), but dropped substantially with more aggressive frame removal (e.g., *τ* = 0.4; ∼ 73% removed; 75% accuracy). More broadly, the results highlight that “highlight” selection must balance time savings against information loss. Moreover, overly aggressive thresholding can eliminate transitional context and reduce temporal coherence, which may be important for recurrent models. This highlights-reel paradigm is particularly relevant to real-world implementation, where manual video-based skill assessment and feedback can be time- and resource-intensive. Even modest reductions in review time could make structured assessment workflows more scalable while preserving clinician oversight, aligning with broader directions in surgical data science emphasizing workflow integration and usable, human-centered AI systems [26].

In contrast to two-stream action recognition paradigms [21], the optical flow variant showed markedly reduced performance on this setting (Table 1). A likely explanation is that optical flow in endoscopic video can be corrupted by camera motion, zoom, specularities, smoke, and nonrigid tissue deformation, producing high-magnitude motion unrelated to technical dexterity. In addition, JIGSAWS trials may contain motion patterns that are not sufficiently stationary across subjects and repetitions, limiting the discriminative value of raw flow fields. These findings suggest that if motion cues are to be incorporated, they may require stabilization or camera-motion compensation, more robust motion representations (e.g., learned flow, residual motion after stabilization), or explicit tool- or region-conditioned motion features rather than whole-frame flow.

Several limitations should be considered. First, JIGSAWS is a controlled, benchtop robotic training dataset and may not fully represent the variability of real-world laparoscopic surgery (occlusion, bleeding, smoke, variable anatomy, changing camera viewpoints, and multi-actor teams). Second, saliency methods such as Grad-CAM provide coarse localization and can sometimes be sensitive to model architecture and training conditions. Thus, explanations should be interpreted as supportive evidence rather than definitive causal attribution. Third, the highlights-reel approach used frame-wise thresholding, which can disrupt temporal continuity. Future implementations may benefit from segment-based selection (contiguous high-saliency intervals) to preserve narrative flow and facilitate human review. Finally, the current evaluation focuses on internal validity within JIGSAWS while external validation on clinical datasets is essential before deployment.

Next steps include validating this pipeline on real operative laparoscopic videos (e.g., inguinal hernia repair), where assessment tools and outcomes may differ from robotic bench tasks. Methodologically, future work should explore segment-level highlight generation with temporal smoothing to improve continuity, learn thresholds adaptively per procedure or per surgeon to maintain a desired accuracy-compression target, and perform a clinician study comparing expert judgments using full videos versus AI-generated highlights to test non-inferiority. These efforts will help determine whether interpretable AI-driven highlights can provide faster, clinically acceptable competency assessment while preserving surgeon trust and oversight.

## 5. Conclusion

We presented an interpretable, video-only deep learning pipeline for automated surgical skill assessment and the generation of AI-driven highlights reels. On the JIGSAWS benchmark, our InceptionV3+GRU RGB model achieved 95% accuracy for three-class expertise classification, with spatial and temporal saliency maps providing clinically plausible explanations. By applying temporal-saliency thresholding, we enabled the construction of shortened surgical highlights and quantified the explicit accuracy-compression trade-off, demonstrating that moderate compression maintains reliability while aggressive thresholding degrades performance. Future work will validate this approach on clinical datasets, specifically laparoscopic inguinal hernia repair, and conduct a non-inferiority study comparing expert assessment of AI-generated highlights against full-length video review.

## Data Availability

The datasets analyzed during the current study (JIGSAWS) are publicly available. The code for the inference pipeline and the trained models produced in this work will be made openly available on GitHub and Hugging Face.

https://cirl.lcsr.jhu.edu/research/hmm/datasets/jigsaws_release/

## Acknowledgments

We acknowledge the Surgical Performance Enhancement and Robotics (SuPER) Centre and the McGill University Health Centre (MUHC) for institutional support. This work was supported in part by the Montreal General Hospital Foundation (MGHF) through the Mimi Dupuis Benjamin Award and the Nesbitt-McMaster Award.

## Author Contributions

M.L. conceived the study, developed the methodology and software, conducted experiments and analysis, and drafted the manuscript. L.S.F. and A.H. supervised the work and revised the manuscript.

## Conflict of Interest

The authors declare that they have no conflict of interest.

## Ethics Approval and Consent to Participate

This study used the publicly available JIGSAWS dataset [13]. No patient data were collected or analyzed for this work.

## Notes

### Competing Interest Statement

The authors have declared no competing interest.

### Author Declarations

The Research Ethics Board of the McGill University Health Centre waived ethical approval for this work. This study exclusively utilized the existing, de-identified JIGSAWS dataset. No new patient data were collected or analyzed for this work.

